# Effects of Dehydroepiandrosterone in Pulmonary Hypertension (EDIPHY): A Randomized, Double-Blind, Placebo-Controlled Crossover Trial

**DOI:** 10.1101/2025.08.01.25332627

**Authors:** Rachel Sanders, Thomas Walsh, Grayson L. Baird, Michael K. Atalay, Saurabh Agarwal, Daniel Arcuri, James R. Klinger, Christopher J. Mullin, James Simmons, Navneet Singh, Mary Whittenhall, Corey E. Ventetuolo

## Abstract

**Rationale:** The adrenal steroid dehydroepiandrosterone (DHEA) and its sulfated form (DHEA-S) are deficient in pulmonary arterial hypertension (PAH) and lower levels are associated with worse right ventricular (RV) function, a decisive factor for survival in PAH.

**Objectives:** We sought to determine whether DHEA improved RV function as measured by cardiac magnetic resonance imaging (MRI) in PAH.

**Methods:** We conducted a randomized, double-blind, placebo-controlled crossover clinical trial of DHEA in participants with PAH at a single center. All participants and study staff were blinded. The primary outcome was change in RV longitudinal strain as measured by MRI after 18 weeks. Key secondary outcomes were the change in serum DHEA-S levels and other PAH end points after 18 weeks.

**Measurements and Main Results:** A total of 26 participants were randomized to DHEA first or placebo first between 2019 and 2024, 20 (77%) of whom were females. Twenty-three (88%) participants completed the study; one participant completely withdrew. DHEA had no effect on RV longitudinal strain. DHEA improved RV short axis radial strain (20.7% [95% CI 16.7, 24.6] to 23.1% [95% CI 19.5, 26.7] compared to placebo (21.1% [95% CI 18.1, 24.0] to 19.3% [95% CI 16.5, 22.2])(p = 0.031), as well as emPHasis-10 scores (p = 0.037) and Short Form-36 physical component scores (p = 0.044) in the second treatment period. In both treatment periods, DHEA may have worsened RV circumferential strain as compared to placebo (p = 0.047, p = 0.052 respectively). Active treatment with DHEA significantly increased serum DHEA-S levels at the end of both treatment periods (p<0.0001), which was in turn associated with multiple study end points in the predicted direction (higher DHEA-S levels, improved PAH metrics). Active treatment with DHEA significantly increased serum testosterone levels as compared to placebo (p <0.01 for both treatment periods), which may have counteracted effects of DHEA supplementation. There was no difference in adverse events.

**Conclusions:** DHEA treatment did not improve RV longitudinal strain in this pilot crossover trial but had variable and possibly beneficial effects on other PAH endpoints. DHEA increased DHEA-S and testosterone levels, which may explain some of the discordant results. DHEA is safe and well tolerated in PAH.

**Clinical trial registered with** www.clinicaltrials.gov **(NCT03648385)**

## Introduction

Biological sex and sex hormones are known to modulate pulmonary arterial hypertension (PAH) and right ventricular (RV) adaptation, the major determinant of outcome in PAH. In studies using both supervised and unsupervised approaches, circulating levels of the adrenal steroid dehydroepiandrosterone (DHEA) and its sulfate ester (DHEA-S) are lower in PAH as compared to healthy controls and associated with worse outcomes including RV dysfunction, independent of other sex hormones, biological sex, and menopausal status (1–5). These human data complement preclinical models demonstrating that DHEA reverses pulmonary hypertension (PH) and rescues the RV by reducing oxidative stress (6–10). Further, DHEA binds directly to vascular endothelium to activate nitric oxide synthase, promotes pulmonary vascular relaxation, suppresses endothelin-1 expression, and inhibits cardiac remodeling (11, 12).

In addition to being available as an over-the-counter dietary supplement, DHEA has been studied in randomized controlled trials (RCT) in patients with adrenal insufficiency and connective tissue disease, with mixed results (13–23). In a single arm study of eight patients with PH related to chronic obstructive pulmonary disease (COPD), treatment with three months of DHEA was associated with a significant increase in six-minute walk distance (6MWD) and improvements in pulmonary hemodynamics without adverse effects (24). Thus, since PAH patients have low levels of DHEA-S that are associated with worse disease metrics including RV dysfunction, and because exogenously administered DHEA improves oxidative signaling and may be beneficial in related conditions, a clinical trial to examine the safety and efficacy of DHEA in PAH was warranted.

We conducted a forty-two-week single center crossover trial of DHEA supplementation in participants with PAH. We hypothesized that DHEA supplementation would raise circulating levels of DHEA-S in participants and improve RV structure and function as measured by cardiac magnetic resonance imaging (MRI) as compared to placebo in patients with PAH.

## Methods

### Trial Design

The Effects of Dehydroepiandrosterone in Pulmonary Hypertension (EDIPHY) trial was a randomized, double-blind, placebo-controlled crossover trial of DHEA in females and males with PAH funded by the National Heart, Lung, and Blood Institute. The study design has previously been published (25); the research protocol was approved by the Rhode Island Hospital Institutional Review Board (IRB) (IRB #001218). Study conduct and subject safety was monitored by a Data Safety and Monitoring Board. The trial was registered on clinicaltrials.gov (NCT03648385) and received an Investigational New Drug exemption (IND #129285) from the U.S. Food and Drug Administration before recruitment began.

### Study Participants

Participants were recruited from the Brown University Health Pulmonary Hypertension Center and regional PH support networks. Patients with PAH and a history of classical hemodynamic criteria documented by right heart catheterization (mean pulmonary artery pressure ≥ 25 mmHg; pulmonary artery wedge pressure ≤ 15 mmHg; pulmonary vascular resistance > 3 Wood units) who were able to undergo MRI were included (26). We included subjects older than 18 years of age and females regardless of menopausal status. We excluded PAH associated with human immunodeficiency virus (27). Patients on active systemic hormonal therapy, contraceptives, or supplements (or considering their use) were excluded while hormone-containing intrauterine devices were permissible (28). Patients with an established history of non-adherence or other circumstances which could threaten adherence with the crossover design and visit schedule were not approached.

Treated PAH patients were eligible provided they had no new PAH therapies introduced for 12 weeks and were clinically stable (no hospitalizations or up-titration of PAH therapies) for four weeks between screening and randomization. A detailed list of inclusion and exclusion criteria is included in Table E1 of the online supplement. All participants provided written informed consent.

### Study Procedures

The protocol compared over-encapsulated DHEA 50 mg daily (Green Mountain Pharmaceuticals; Lakewood, CO) versus identical placebo in two 18-week treatment periods (Periods 1 and 2) which included five study visits, three MRIs and a four-week washout period (Figure 1). Subjects were randomized to placebo or DHEA (Period 1) and then switched (DHEA vs placebo) in Period 2 and stratified by sex. Highly pure DHEA (>96.5%) was verified by an independent laboratory (Infinity Laboratories, Castle Rock, CO). Participants, study investigators, study staff and the study statistician were all masked. Following screening and informed consent, a maximum of four weeks were allowed to lapse before randomization at the baseline visit to confirm clinical stability and that there were no recent changes to PAH medications.

**Figure 1.**
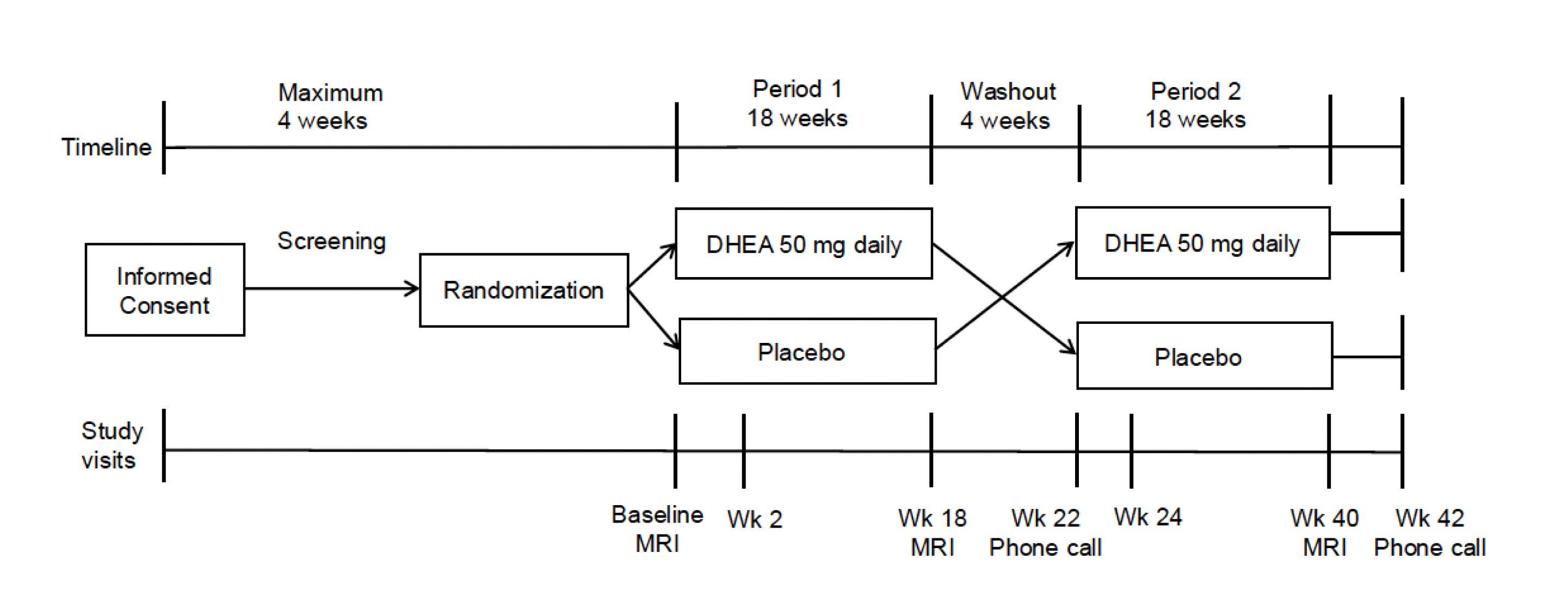
Study design schematic. Magnetic resonance (MRI) was performed at baseline, week 18 (end of Period 1), and week 40 (end of Period 2). Other end points were also assessed at week 2 and week 24. DHEA=dehydroepiandrosterone.

The protocol called for the enrollment of 26 participants. The primary outcome was change in RV longitudinal strain as measured by cardiac MRI after 18 weeks. A key secondary outcome was the change in serum DHEA-S levels after 18 weeks. Additional endpoints included other RV parameters, World Health Organization functional class, 6MWD, health-related quality of life (emPHasis-10, Short Form-36 (SF-36)), N-terminal pro hormone of brain natriuretic peptide (NT-proBNP) levels, other sex hormone and biomarker levels, side effects and safety. Additional details of study procedures, end point assessments, and monitoring have been previously published and are included in the online supplement (Table E2)(25).

### Statistical Methods

All analyses were conducted using SAS Software 9.4 (Cary, NC). Outcomes were modeled over time (baseline, end of Period 1, end of Period 2). The primary and secondary outcomes were examined using generalized estimating equations (GEE) with sandwich estimation to correct for model misspecification using normal, lognormal, or negative binomial distributions, when appropriate. Sex as a moderator was considered for certain exploratory analyses.

As it is unknowable in a crossover trial whether the washout period is sufficient to avoid carry-over effects and as study end points were collected at a second baseline (week 22), change in all outcomes were examined between 1) treatment (DHEA vs. placebo) and 2) treatment and period. All analyses were conducted according to the intention-to-treat principle. In the case of partial withdrawal (discontinuation of study drug), study assessments were still completed per protocol. Alpha was established a priori at the 0.05 level and all interval estimates were calculated for 95% confidence.

## Results

We assessed 73 patients for eligibility. A total of 26 participants were randomized between January 2019 and July 2024, 20 (77%) of whom were females. Twenty-three (88%) participants completed the study; two participants partially withdrew (i.e., stopped study drug but continued in the study protocol) and one participant completely withdrew (Figure 2). A greater proportion of the participants who were randomized to DHEA in the first treatment period were male (four out of 14) and premenopausal females (six out of 14) compared to participants randomized to DHEA in the second treatment period (two males and two premenopausal females out of 12). Those randomized to DHEA in the first treatment period also tender to have longer 6MWD, more favorable hemodynamics, higher RV ejection fraction (RVEF) and lower RV end-diastolic and end-systolic volumes (RVEDV; RVESV) (Table 1). The two groups (DHEA to placebo and placebo to DHEA) were similar in terms of body mass index and functional class.

**Figure 2.**
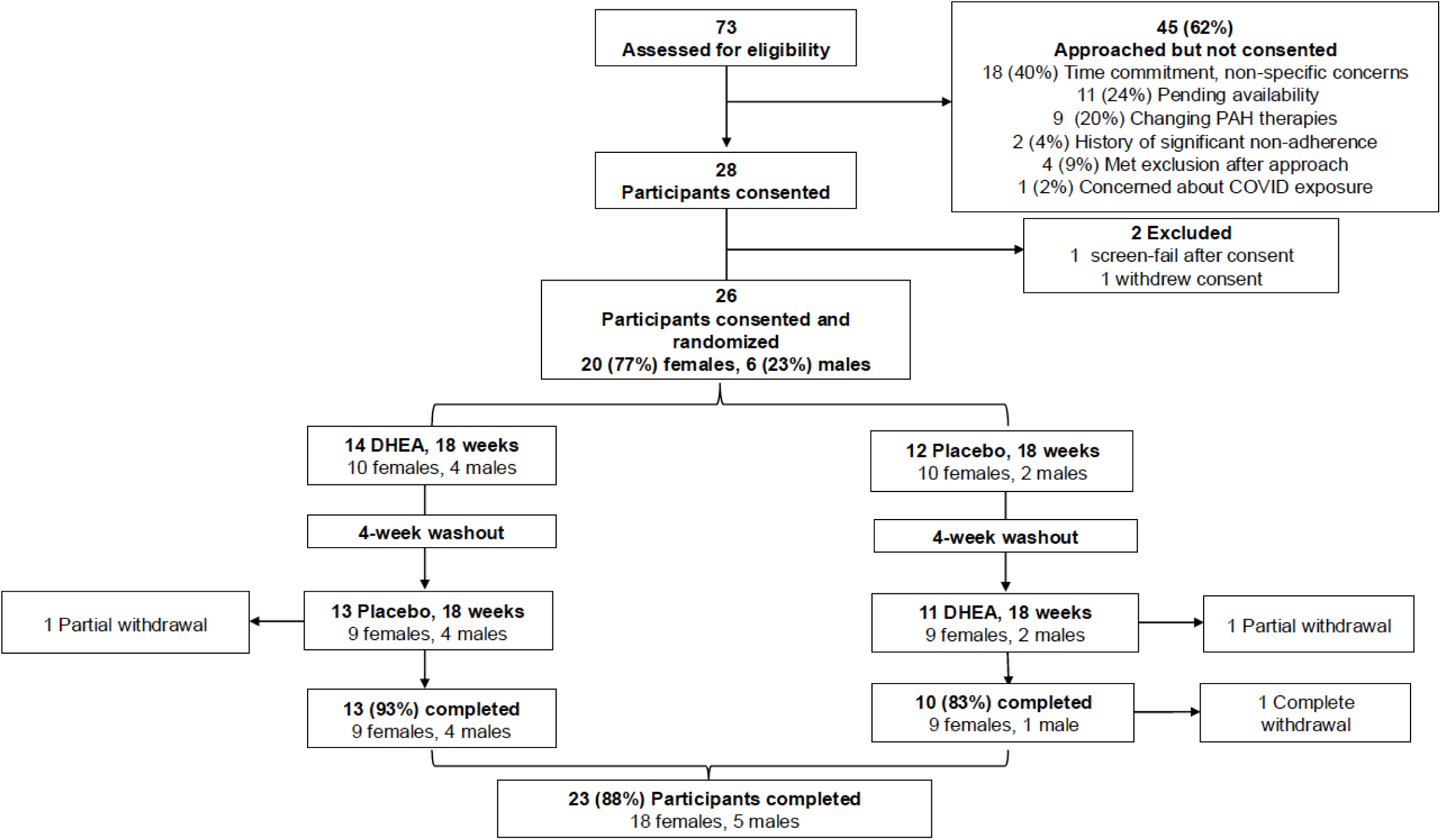
CONSORT (Consolidated Standards of Reporting Trials) diagram. PAH=pulmonary arterial hypertension; DHEA=dehydroepiandrosterone.

**Table 1.** Baseline characteristics of participants randomized to treatment sequence.

| **Variables** | **Treatment sequence** |  |  |
| --- | --- | --- | --- |
|  | **DHEA to placebo** | **Placebo to DHEA** | **Total** |
| Number | 14 | 12 | 26 |
| Male, n (%) | 4 (29) | 2 (17) | 6 (23) |
| Female, n (%) | 10 (71) | 10 (83) | 20 (77) |
| Premenopausal, n (%) | 6 (43) | 2 (17) | 8 (31) |
| Postmenopausal, n (%) | 4 (29) | 8 (67) | 12 (46) |
| Age, yr | 44 (38 – 55) | 54 (46 – 68) | 48 (39 – 69) |
| BMI, kg/m<sup>2</sup> | 32 (28 – 37) | 34 (28 – 36) | 32 (27 – 36) |
| Race, n (%) |  |  |  |
| White | 11 (79) | 9 (75) | 20 (77) |
| Black | . | 1 (8) | 1 (4) |
| Other / Multiple | 4 (29) | 2 (17) | 6 (23) |
| Hispanic ethnicity, n (%) | 3 (21) | 1 (8) | 4 (15) |
| PAH Diagnosis, n (%) |  |  |  |
| Idiopathic | 5 (36) | 5 (42) | 10 (38) |
| Heritable | 3 (21) | 2 (17) | 5 (19) |
| Drug and toxin | . | 1 (8) | 1 (4) |
| Connective tissue disease | 5 (36) | 4 (33) | 9 (35) |
| Congenital heart disease | 1 (7) | . | 1 (4) |
| Functional class, n (%) |  |  |  |
| II | 11 (79) | 8 (67) | 19 (73) |
| III | 3 (21) | 4 (33) | 7 (27) |
| Baseline 6MWD, m | 437 (391 – 477) | 420 (230 – 451) | 430 (352 – 457) |
| N-terminal proBNP, pg/mL | 79.9 (63.3 – 193.8) | 193.2 (104.4 – 297.5) | 127.6 (67.3 – 239.8) |
| Hemodynamics |  |  |  |
| Right atrial pressure, mm Hg | 7 (6 – 12) | 10 (7 – 13) | 10 (7 – 13) |
| Mean PAP, mm Hg | 43 (39 – 52) | 50 (43 – 57) | 45 (34 – 54) |
| PAWP, mm Hg | 11 (8 – 11) | 13 (9 – 13) | 11 (8 – 13) |
| Cardiac index, L/min/m<sup>2</sup> | 2.9 (2.7 – 3.5) | 2.7 (1.8 – 2.8) | 3 (2 – 3) |
| PVR, Wood units | 5 (4 – 8) | 8 (6 – 11) | 6 (5 – 9) |

| Health-Related Quality of Life |  |  |  |
| --- | --- | --- | --- |
| EmPHasis-10 | 25 (20 – 29) | 19 (12 – 29) | 25 (18 – 29) |
| Short-Form 36, PCS | 38 (36 – 42) | 45 (37 – 48) | 40 (36 – 46) |
| Short-Form 36, MCS | 48 (42 – 57) | 54 (46 – 57) | 49 (43 – 57) |
| Baseline MRI measures |  |  |  |
| RV longitudinal strain, % | -18.4 (-20.4, -17.0) | -18.8 (-20.0, -13.5) |  |
| RV peak global radial strain, % | 22.4 (18.6, 26.8) | 18.9 (16.3, 22.3) | 20 (16.9, 25.7) |
| RV circumferential strain, % | -14.8 (-16.8, -12.3) | -12.2 (-14.2, -11.0) | -13.3 (-16.3, -11.2) |
| RVEF, % | 55 (51 – 58) | 51 (44 – 57) | 53 (48 – 57) |
| RVEDV, mL | 147 (123 – 182) | 181 (125 – 201) | 163 (123 – 195) |
| RVESV, mL | 62 (53 – 85) | 92 (55 – 114) | 71 (53 – 113) |
| RV stroke volume, mL | 82 (70 – 95) | 83 (69 – 97) | 82 (68 – 95) |
| RV mass, g | 55 (39 – 88) | 55 (40 – 78) | 55 (39 – 88) |

Data shown as median (interquartile range) or number, %. DHEA=dehydroepiandrosterone; BMI=body mass index; PAH=pulmonary arterial hypertension; 6MWD=six-minute walk distance; PAP=pulmonary artery pressure; PAP=pulmonary artery wedge pressure; PVR=pulmonary vascular resistance; PCS=physical component score; MCS=mental component score; RV=right ventricular; RVEF=RV ejection fraction; RVEDV=RV end-diastolic volume; RVESV= RV end-systolic volume.

### Effects of DHEA on ventricular function

There was no evidence of effect of DHEA on RV longitudinal strain (Figure 3A; Table 2). DHEA improved RV short axis radial strain in the second treatment period (20.7% [95% CI 16.7, 24.6] to 23.1% [95% CI 19.5, 26.7] compared to placebo (21.1% [95% CI 18.1, 24.0] to 19.3% [95% CI 16.5, 22.2])(p = 0.031)(Figure 3B. In both treatment periods, DHEA may have worsened (less negative) RV circumferential strain as compared to placebo (e.g., Period 1: DHEA first -14.3% [95% CI 15.9, -12.6] to -13.7% [95% CI - 15.2, -12.2] vs. Placebo first -12.3 [95% CI -13.9, -10.6] to -13.3 [95% CI -15.5, -11.2]; p = 0.047)(Fig 3C). DHEA increased RV end-diastolic volume over the first treatment period as compared to placebo (156.7 mL [95% CI 133.7, 183.7] to 160.4 mL [95% CI 137.3, 187.4] vs. 173.8 mL [95% CI 140.2, 215.5] to 159.8 mL [95% CI 130.1, 196.3]; p = 0.044). There was no evidence of effect of DHEA on RV ejection fraction, other volumes, or mass over placebo (Table 3). Active treatment with DHEA did not affect left ventricular (LV) measures including LV ejection fraction, volumes and mass (Table E3).

**Figure 3.**
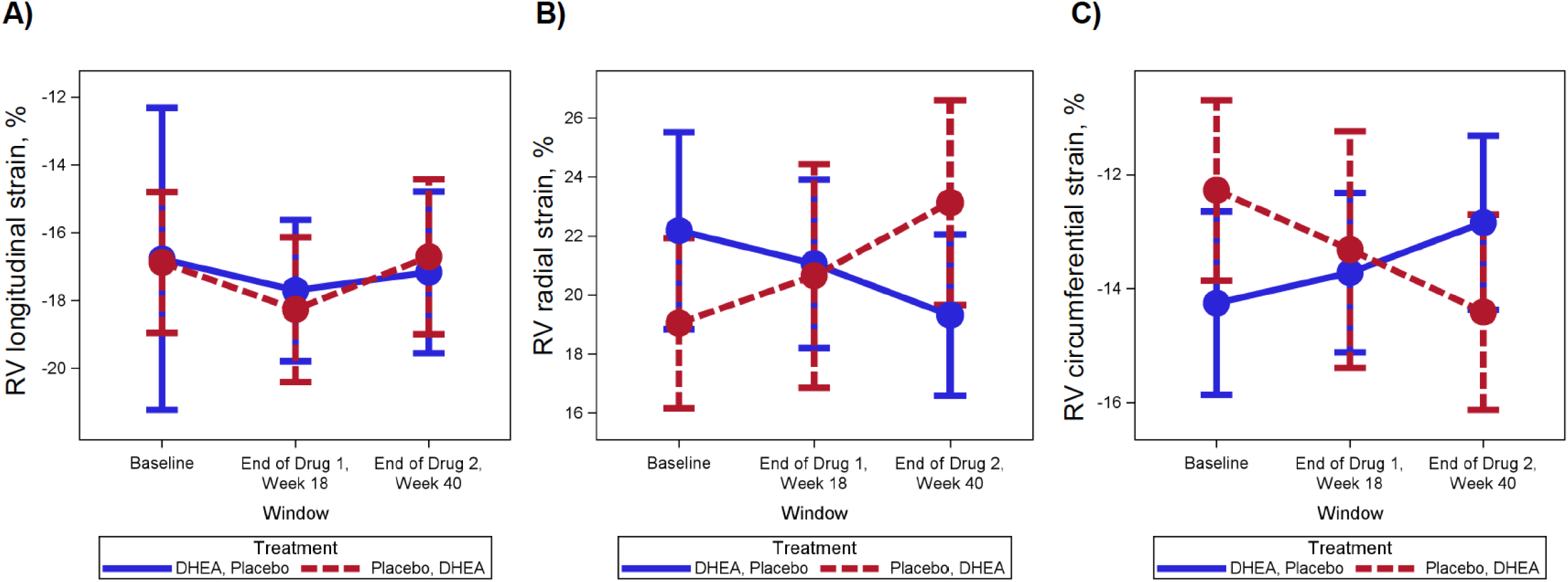
Longitudinal changes in right ventricular (RV) strain measurements across study windows. **A)** RV longitudinal strain. **B)** RV short axis radial strain. **C)** RV short axis circumferential strain. Both panels: Blue=randomized to DHEA first (baseline – week 18) then placebo (week 22 – week 40); Red=randomized to placebo first (baseline – week 18) then DHEA (week 22 – 40).

**Table 2.**
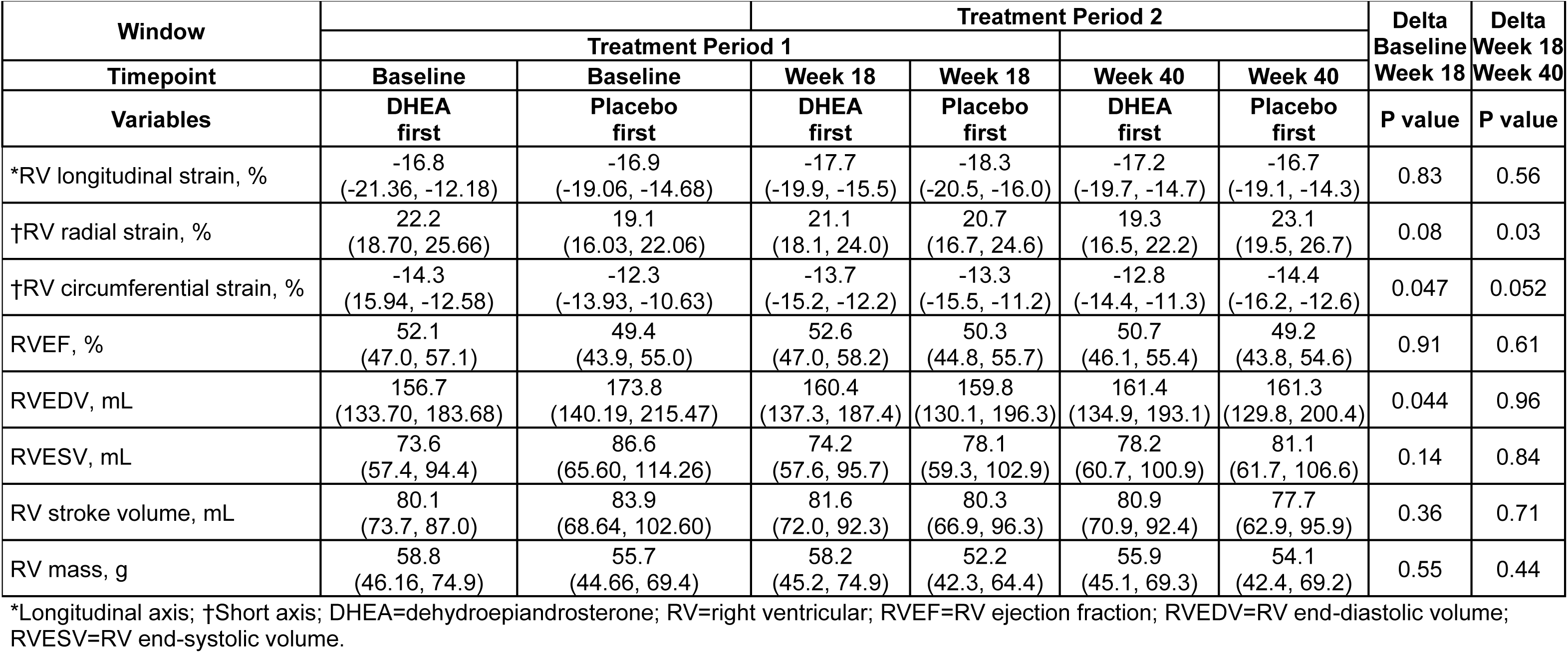
Right ventricular cardiac magnetic resonance imaging measures after DHEA and placebo.

**Table 3.**
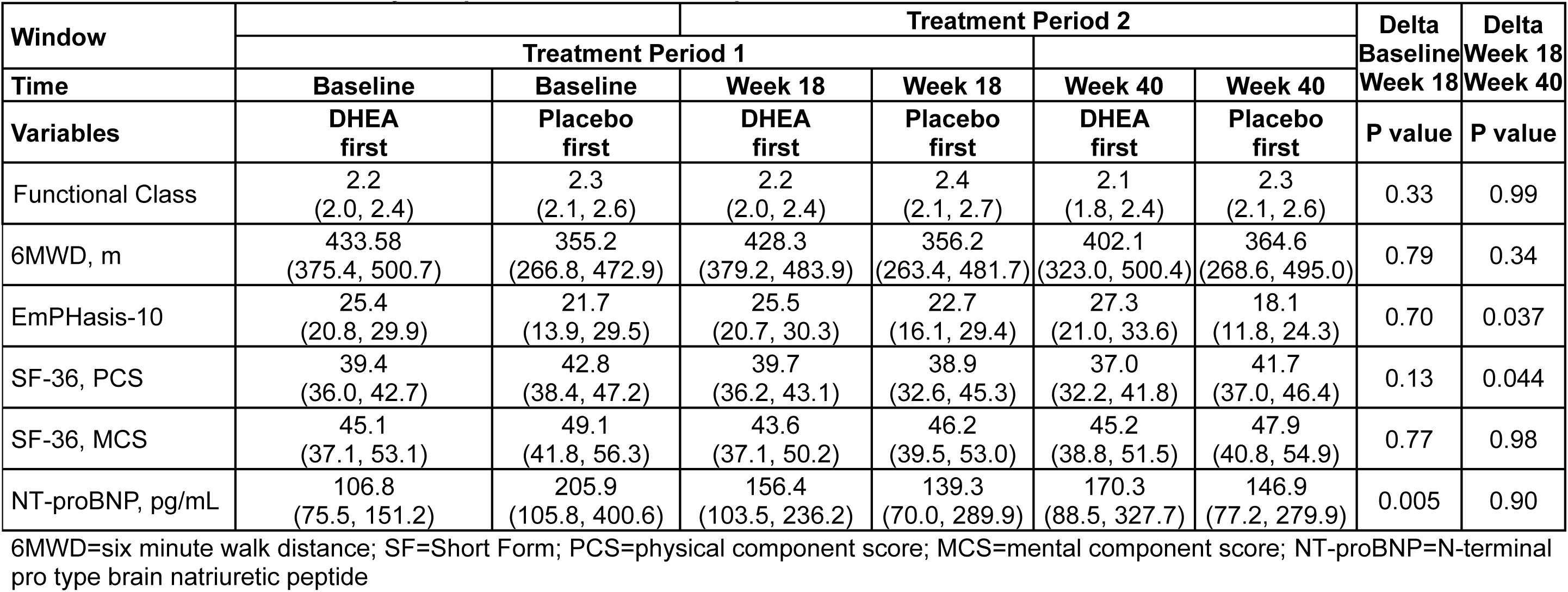
Differences in secondary end points after DHEA and placebo.

### Effects of DHEA on other PAH measures

There was no evidence that DHEA effected functional class or 6MWD over placebo (Table 3). EmPHasis-10 scores improved (lower scores, better HRQoL) in the second treatment period for those on DHEA (22.7 [95% CI 16.1, 29.4] to 18.1 [95% CI 11.8 to 24.3]) compared to placebo (25.5 [95% CI 20.7, 30.3] to 27.3 [95% CI 21.0, 33.6])(p=0.037). DHEA increased physical component score of the SF-36 (higher scores, better HRQoL) in the second treatment period (38.9 [95% CI 32.6, 45.3] to 41.7 [95% 37.0, 46.4] as compared to placebo (39.7 [95% CI 36.2, 43.1] to 37.0 [95% CI 32.2, 41.8](p = 0.044). NT-proBNP levels increased in the first window for those on DHEA (106.8 pg/mL [95% CI 75.5, 151.2] to 156.4 pg/mL [95% CI 103.5, 236.2]) compared to placebo (205.9 pg/mL [95% CI 105.8, 400.6] to 139.3 pg/mL [95% CI 70.0, 289.9](p=0.005).

### DHEA-S levels, randomization sequence and other sex hormone levels

Active treatment with DHEA significantly increased serum DHEA-S levels at the end of both treatment periods (p<0.0001)(Table 4; Figure 4A). Participants who were randomized to DHEA in the first window experienced a greater increase in DHEA-S than participants randomized to DHEA in the second window (77.8 μg/dL [95% CI 49.0, 123.8] to 287.8 μg/dL [95% CI 197.5, 419.3] vs. 49.3 μg/dL [95% CI 28.2, 86.1] to 169.9 μg/dL [95% CI 87.0, 332.0](p = 0.034) (Figure 4B). Biological sex may have moderated these changes (Figure 4C), as males had higher DHEA-S levels than females (especially females in the placebo first group)(p=.007). Those that were randomized to receive DHEA first had a decline in their levels which began while still on active treatment (Week 18, Visit 3) and returned to baseline after the crossover, two weeks into Period 2 (Week 24, Visit 4) and through the end of the study period (Week 40).

**Figure 4.**
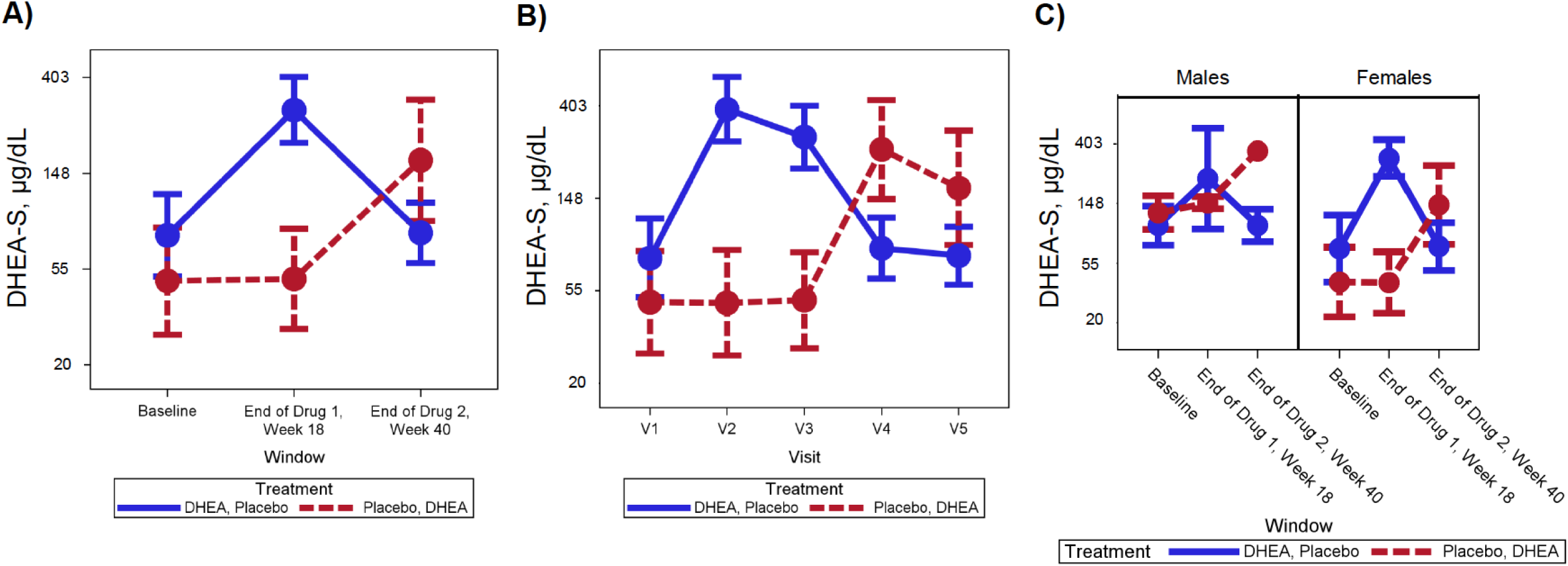
A) Longitudinal changes in dehydoepiandrosterone-sulfate (DHEA-S) levels across study windows by randomization sequence. **B)** Longitudinal changes in DHEA-S levels across study visits (V1 – V5). **C)** Longitudinal changes in DHEA-S levels across study windows by randomization sequence and sex. All panels: Blue=randomized to DHEA first (baseline – week 18) then placebo (week 22 – week 40); Red=randomized to placebo first (baseline – week 18) then DHEA (week 22 – 40).

**Table 4.** Differences in hormones and other biomarkers after DHEA and placebo.

| Window |  |  | Treatment Period 2 |  |  |  | Delta | Delta |
| --- | --- | --- | --- | --- | --- | --- | --- | --- |
|  | Treatment Period 1 |  |  |  |  |  | Baseline | Week 18 |
| Time | Baseline | Baseline | Week 18 | Week 18 | Week 40 | Week 40 | Week 18 | Week 40 |
| Variables | DHEA first | Placebo first | DHEA first | Placebo first | DHEA first | Placebo first | P value | P value |
| DHEA-S, µg/dL | 77.8<br>(49.0, 123.8) | 48.2<br>(26.6, 87.5) | 287.8<br>(197.5, 419.3) | 49.3<br>(28.2, 86.1) | 75.9<br>(55.0, 104.6) | 169.9<br>(87.0, 332.0) | <0.0001 | <0.0001 |
| Estradiol, pg/mL | 46.3<br>(22.9, 93.6) | 26.3<br>(13.9, 49.7) | 60.7<br>(33.0, 111.7) | 24.7<br>(11.2, 54.3) | 34.5<br>(18.0, 66.3) | 25.3<br>(14.3, 44.8) | 0.36 | 0.23 |
| Testosterone, ng/dL | 23.8<br>(9.0, 62.7) | 10.2<br>(3.8, 27.3) | 65.6<br>(37.3, 115.4) | 10.4<br>(3.7, 29.3) | 27.0<br>(10.8, 67.4) | 35.3<br>(14.2, 87.7) | 0.002 | <0.0001 |
| Progesterone, ng/mL | 0.6<br>(0.4, 1.0) | 0.6<br>(0.3, 1.3) | 0.6<br>(0.3, 0.9) | 0.4<br>(0.3, 0.6) | 0.8<br>(0.4, 1.6) | 0.4<br>(0.2, 0.6) | 0.42 | 0.28 |
| FSH, mIU/mL | 11.3<br>(6.7, 18.9) | 21.5<br>(11.8, 39.1) | 9.7<br>(5.7, 16.6) | 23.5<br>(13.0, 42.7) | 9.4<br>(5.4, 16.7) | 25.5<br>(14.7, 44.2) | 0.08 | 0.65 |
| SHBG, nmol/L | 57.8<br>(41.7, 79.9) | 61.0<br>(44.2, 84.21) | 50.2<br>(38.1, 66.1) | 62.2<br>(44.2, 87.4) | 59.3<br>(43.1, 81.6) | 48.0<br>(34.2, 67.5) | 0.07 | <0.01 |
| Luteinizing hormone, mIU/mL | 12.5<br>(7.9, 12.0) | 14.3<br>(8.4, 24.3) | 10.3<br>(6.3, 16.7) | 14.5<br>(8.3, 25.4) | 8.6<br>(4.7, 15.6) | 17.5<br>(11.8, 26.1) | 0.27 | 0.26 |
| Prolactin, µIU/mL | 329.5<br>(274.8, 395.1) | 287.8<br>(206.4, 401.1) | 327.4<br>(256.2, 418.4) | 293.8<br>(212.6, 406.1) | 305.2<br>(267.3, 348.5) | 271.4<br>(190.1, 387.6) | 0.81 | 0.94 |
| C-peptide, ng/mL | 3.3<br>(2.5, 4.5) | 3.6<br>(2.5, 5.3) | 3.1<br>(2.3, 4.1) | 3.3<br>(2.3, 4.7) | 3.0<br>(2.3, 4.0) | 3.4<br>(2.5, 4.6) | 0.93 | 0.83 |
| Insulin, µU/mL | 11.4<br>(7.5, 17.4) | 13.2<br>(7.5, 23.4) | 10.7<br>(6.7, 17.0) | 12.1<br>(7.2, 20.2) | 10.8<br>(7.1, 16.4) | 13.5<br>(8.0, 22.8) | 0.85 | 0.74 |
| Cortisol, µg/dL<sup>2</sup> | 11.7<br>(8.9, 15.4) | 9.1<br>(6.8,12.1) | 9.2<br>(6.7, 12.8) | 12.6<br>(10.3, 15.4) | 12.9<br>(9.9, 16.6) | 11.7<br>(9.7, 14.4) | 0.004 | 0.05 |

DHEA-S=dehydroepiandrosterone-sulfate; FSH=follicle stimulating hormone; SHBG=sex hormone binding globulin

Similarly, those who received DHEA second had a decline in their levels over the second treatment period.

Active treatment with DHEA significantly increased serum testosterone levels (23.8 to 65.6 ng/dL) compared to placebo (10.2 to 10.4 ng/dL) during the first treatment period (p=0.002) and the second treatment period (10.4 to 35.3 ng/dL vs. 65.6 to 27.0 ng/dL) (p<0.0001). DHEA decreased sex hormone binding globulin over Period 1 (p = 0.07) and Period 2 (p < 0.01), and decreased cortisol levels (Period 1: p = 0.004; Period 2: p = 0.05) as compared to placebo.

### Associations between changes in DHEA-S levels and end points

We observed multiple relationships between serum DHEA-S levels and study end points, in the hypothesized direction. Higher levels of DHEA-S may have been associated with improved RV radial strain (p = 0.063) (Figure E1A). Similarly, higher levels of DHEA-S may have been associated with better (more negative) circumferential strain (p = 0.080) (Figure E1B) as well as possibly lower NT-proBNP levels (p = 0.138)(Figure E2A). Higher levels of DHEA-S appeared associated with longer 6MWD (p = 0.060) (Figure E2D), higher physical component scores of the SF-36 (p = 0.007) and lower emPHasis-10 scores (p = 0.126).

### Interaction with testosterone levels

While DHEA increased DHEA-S levels, we did not observe consistent associations between active treatment-DHEA-S levels and improved outcomes. This may be explained by the observed increases in testosterone, as DHEA is a prohormone for testosterone and testosterone has been shown to have detrimental effects on the RV (29). As is shown in the 3-dimensional plot (Figure E3), changes in RV longitudinal strain were neutralized as DHEA-S levels and testosterone levels were increased.

### Side Effects and Safety

Adverse events (AEs) occurred in 23 (88%) participants with serious adverse events in 10 (43%). There was no evidence that AEs occurred more on DHEA (average count AEs during active treatment 1.6 vs. placebo 2.5, p=0.61). More AEs occurred during treatment with placebo (n = 53; 50.5% of all AEs including the washout period) than with DHEA (n = 37; 35.2% of all AEs including the washout period). We specifically queried participants about androgenic side effects and skin and nail changes that were to be expected with DHEA. The online supplement includes specific information on side effects and adverse events (Table E4).

## Discussion

This is the first RCT to investigate supplementation of a naturally occurring hormone in PAH and among the first to use RV strain and other RV parameters from MRI as pivotal end points. DHEA did not impact RV longitudinal strain, had variable effects on other MRI measures (improved radial strain, worsened circumferential strain and increased RVEDV), and improved emPHasis-10 and physical component scores of the SF-36, particularly in the second treatment period. Active treatment significantly increased DHEA-S levels, which were in-turn associated with an improvement in some RV parameters, HRQoL as well as 6MWD. DHEA significantly increased testosterone levels (and decreased cortisol levels) which may explain some of the discordant results. DHEA was well tolerated and safe.

While RV strain assessed by MRI is felt to be a highly sensitive measure that may precede worsening of established parameters in PAH, temporal changes over time, definitive directionality and integration of all strain values (globally improved or worsened RV function), as well as known minimally clinically important differences are less well (or not) established in PAH (30, 31). The significant changes in additional study endpoints noted (higher NT-proBNP and RVEDV, improved HRQoL) were relatively small and below established thresholds for risk or minimally important differences in PAH (32–34).

We do not know if longer treatment with DHEA might have demonstrated definitive benefit, or if certain subgroups of patients (i.e., based on sex or pre- vs. post- menopausal status) are more likely to benefit.

In fact, there were baseline imbalances such that those randomized to DHEA first tended to be male or premenopausal females and had more favorable PAH measures and RV function. While male sex is associated with worse hemodynamics and prognosis in PAH (35, 36), on balance, as healthier individuals received DHEA first this may have biased results towards the null, making it less likely to demonstrate DHEA’s benefits (i.e., a ceiling effect). We were not adequately powered to detect benefit during the first treatment period, especially given the relative overrepresentation of males and premenopausal females despite randomization.

Although we successfully increased DHEA-S levels with active treatment, active treatment was not consistently associated with improved outcomes despite levels of DHEA-S tracking directionally with these measures in the hypothesized direction (in line with prior studies by us and others)(1, 2, 4, 5, 36, 37). It is possible that DHEA does not directly lie in the PAH-RV failure causal pathway but does correlate highly with established and major pathobiological mechanisms of disease. Alternatively, we speculate that the effect of DHEA on outcomes could have been counteracted. In male rats exposed to chronic hypoxia, DHEA prevents and reverses pulmonary hypertensive changes, while testosterone has no effect (38). In the RV, both endogenous and exogenous testosterone increases fibrosis and maladaptive responses to load stress in male mice after pulmonary artery banding (29). While DHEA may naturally improve outcomes, when DHEA is increased therapeutically, it raises testosterone, thereby canceling some of DHEA’s salubrious effects. As well, some of the noted improvements (e.g. in HRQoL) may have been due to decreased cortisol levels.

Those who received DHEA second achieved lesser increases in their DHEA-S levels as compared to those reached in the first treatment period, which may be explained by differences in baseline characteristics (although DHEA-S levels were similar at baseline), underlying disease progression, as low circulating DHEA-S levels tracks with more severe disease in PAH (1–5), or degradation of adherence over the course of the trial. Levels achieved over both treatment periods far exceeded those reported for age-, sex- and BMI healthy controls in our prior studies (1, 2, 5), arguing against lack of consistent and sustained increases in DHEA-S as an explanation for the null results; supranormal levels could also explain the complexity of our findings.

There are limitations to this single center, pilot trial. The study design did not have two “true” baselines; e.g., we incorporated three (not four) MRIs to minimize burden and enhance feasibility. This proved to be relevant, in that two patients approached had contraindications to MRI and could not be enrolled, 18/45 (40%) of patients approached declined enrollment because of nonspecific concerns about the study protocol, parenteral therapy patients required a change in their infusion pumps and tubing to be MRI-compatible, and some participants required benzodiazepine treatment during MRIs for claustrophobia. The absence of a second “true” baseline (week 22 was a phone call only) makes it difficult to confirm there were no carry over effects and may have introduced an order effect. While the 50 mg dose of DHEA was lower than the prior open-label trial of 200 mg of DHEA in patients with COPD-PH (24), we confirmed a highly pure compound and successfully increased DHEA-S levels, although we cannot be sure a higher dose would not have resulting in more durable changes or consistent results.

## Conclusion

DHEA treatment did not improve RV longitudinal strain in this pilot crossover trial but had variable and possibly beneficial effects on other PAH endpoints. DHEA increased DHEA-S and testosterone levels and decreased cortisol levels, which may explain some of the discordant results, along with baseline differences in biological sex and menopausal status despite randomization. DHEA is safe and well tolerated in PAH. Taken together, this trial demonstrates the need for ongoing query of sexual dimorphism and the role of sex hormones in RV adaptation in pulmonary vascular disease.

## Data Availability

The results of this clinical trial are being posted on clinicaltrials.gov A metafile will also be available upon publication

## Acknowledgements

We wish to acknowledge and thank all EDIPHY participants.

## Financial conflicts of interest

R.S., T.W., G.L.B., M.K.A, S.A., D.A., J.S., S.S., M.W., J.R.K., N.S. report no disclosures.

C.J.M.: has received personal fees from Merck & Co. and Regeneron, outside of the submitted work. His institution has received fees for the conduct of clinical trials from Merck & Co., Pulmovant, Tenax and United Therapeutics

C.E.V.: has received personal fees from Merck & Co., Janssen Pharmaceuticals and Regeneron, outside of the submitted work. Her institution has received fees for the conduct of clinical trials from Merck & Co., Pulmovant, Tenax and United Therapeutics.

## Notes

### Clinical Trial

NCT03648385

### Funding Statement

This work was completed with support from the National Institutes of Health R01-HL141268 (CEV); 3U54GM115677-09S3 (CEV); R01-HL174007 (CEV)

### Author Declarations

The study design has previously been published; the research protocol was approved by the Rhode Island Hospital Institutional Review Board (IRB) (IRB #001218). Study conduct and subject safety was monitored by a Data Safety and Monitoring Board. The trial was registered on clinicaltrials.gov (NCT03648385) and received an Investigational New Drug exemption (IND #129285) from the U.S. Food and Drug Administration before recruitment began.

